# Accurately Estimating Unreported Infections using Information Theory

**DOI:** 10.1101/2021.09.14.21263467

**Authors:** Jiaming Cui, Bijaya Adhikari, Arash Haddadan, A S M Ahsan-Ul Haque, Jilles Vreeken, Anil Vullikanti, B. Aditya Prakash

**Affiliations:** College of Computing, Georgia Institute of Technology, Atlanta, GA 30332, US; Department of Computer Science, Virginia Tech, Blacksburg, VA 24060, US; Department of Computer Science, The University of Iowa, Iowa City, IA 52242, US; Biocomplexity Institute, University of Virginia, Charlottesville, VA 22904, US; CISPA Helmholtz Center for Information Security, Saarbrücken 66123, Germany; Department of Computer Science, University of Virginia, Charlottesville, VA 22904, US

## Abstract

One of the most significant challenges in combating against the spread of infectious diseases was the difficulty in estimating the true magnitude of infections. Unreported infections could drive up disease spread, making it very hard to accurately estimate the infectivity of the pathogen, therewith hampering our ability to react effectively. Despite the use of surveillance-based methods such as serological studies, identifying the true magnitude is still challenging. This paper proposes an information theoretic approach for accurately estimating the number of total infections. Our approach is built on top of Ordinary Differential Equations (ODE) based models, which are commonly used in epidemiology and for estimating such infections. We show how we can help such models to better compute the number of total infections and identify the parametrization by which we need the fewest bits to describe the observed dynamics of reported infections. Our experiments on COVID-19 spread show that our approach leads to not only substantially better estimates of the number of total infections but also better forecasts of infections than standard model calibration based methods. We additionally show how our learned parametrization helps in modeling more accurate what-if scenarios with non-pharmaceutical interventions. Our approach provides a general method for improving epidemic modeling which is applicable broadly.

## 1 Introduction

One of the most significant challenges in combating against the spread of infectious diseases in population is estimating the number of total infections. Our inability in estimating unreported infections allows them to drive up disease transmission. For example, in the COVID-19 pandemic, a significant number of COVID-19 infections were unreported, due to various factors such as the lack of testing and asymptomatic infections [8, 6, 36, 34, 23]. There were only 23 reported infections in five major U.S. cities by March 1, 2020, but it has been estimated that there were in fact more than 28,000 total infections by then [4], and spread the COVID-19 to the whole US. Similar trends were observed in other countries, such as in Italy, Germany, and the UK [38].

In fact, an accurate estimation of the number of total infections is a fundamental epidemiological question and critical for pandemic planning and response. Therefore, epidemiologists use *reported rate* (*α*_reported_) to capture total infections, which is defined as the ratio of reported infections to total infections [27]. One of the benefits of using this definition is that it includes asymptomatic infections, which may also contribute substantially to spread [37, 24]. To estimate the reported rate, data scientists and epidemiologists have devoted much time and effort to using epidemiological models. There are many carefully constructed Ordinary Differential Equation (ODE) based models that capture the transmission dynamics of different infectious diseases [23, 32, 7, 28, 21, 22, 39, 15, 19, 40, 41, 11, 25]. However, these models still suffer from estimating accurate reported rates, leading to suboptimal total infections estimation. For example, as shown in Figure 1, the Minneapolis Metro Area had only 16 COVID-19 reported infections by March 11, 2020. Although epidemiologists estimate that there were 182 total infections (light green part in the iceberg) using epi models, later studies revealed that there were actually around 300 total infections (iceberg below the sea level) [16, 1, **?**]

**Figure 1.**
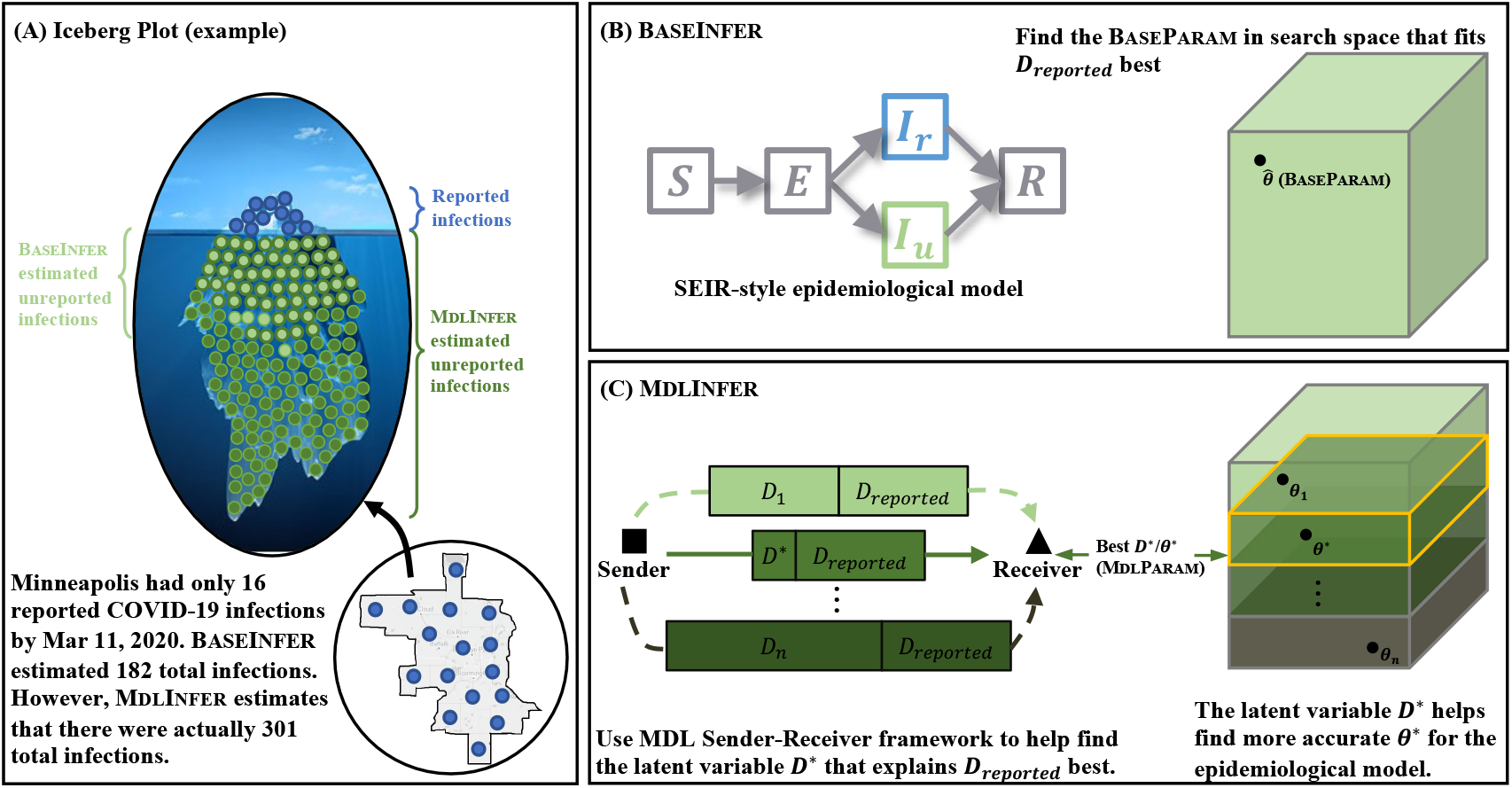
Overview of our problem and methodology. (**A**) We visualize the idea of reported rates using the iceberg. The visible portion above water are the reported infections, which is only a fraction of the whole iceberg representing total infections. Light green corresponds to the 182 unreported infections estimated by typical current practice used by researchers. We call it as the basic approach, or BaseInfer. In contrast, dark green corresponds to the more accurate and much larger 301 unreported infections found by our approach MdlInfer. (**B**) The usual practice is to calibrate an epidemiological model to reported data and compute the reported rate from the resultant parameterization of the model. Here, an SEIR-style model with explicit compartments for reported-vs-unreported infection is shown in the figure as an example. (**C**) Our new approach MdlInfer instead aims to compute a more accurate reported rate by finding a ‘best’ parametrization *for the same epidemiological model* (i.e., SEIR-style model in this example) using a principled information theoretic formulation - two-part ‘sender-receiver’ framework. Assume that a hypothetical Sender *S* wants to transmit the reported infections as the Data to a Receiver *R* in the cheapest way possible. Hence *S* will find/solve for the best *D*\*, intuitively, the Model that takes the fewest number of bits to encode the Data. Using *D*\*, we can find the best Θ*. by exploring a smaller search space.

To tackle this, we propose a new information theory-based approach named MdlInfer to estimate the reported rate. It is based on the following central intuition: Suppose an “oracle” gives us the time series of the number of *total infections D*, we should be able to describe *D*_reported_ in a succinct way: As we know *D*, it is trivial to get the reported rate *α*_reported_. Then with both *D* and *α*_reported_, it will be trivial to describe *D*_reported_, as it is simply *D × α*_reported_ plus a little bit of noise. In practice, we are of course not given *D*, but we could estimate *D* as a latent variable. Specifically, as shown in Figure 1(C), we use Minimum Description Length (MDL) principle to estimate *D*, which allows us to most succinctly describe (i.e., most accurately encode/reconstruct) the dynamics of *D*_reported_. Here, MdlInfer gives an estimate of 301 total infections in Minneapolis as shown below the sea level, which is much closer to the ground truth total infections numbers of around 300 [16, 1].

Our main contributions are summarized below.

- We propose an MDL-based approach on top of ODE-based epidemiological models, which are harder to formulate and optimize. To the best of our knowledge, we are the first to propose an MDL-based approach on top of ODE-based epidemiological models.
- Our proposed MDL-based approach MdlInfer performs superior to the state of the art epidemiological model in estimating total infections and predicting the future reported infections.
- We also show that MdlInfer can aid policy making by analyzing counter-factual non-pharmaceutical interventions, while inaccurate epidemiological model estimates may lead to wrong non-pharmaceutical intervention conclusions.

The rest of the paper is organized in the following way: Section 2 discusses the related works. In section 3, we introduce the current ODE model calibration method to estimate the reported rate, and the background of MDL framework. We then introduce our MdlInfer framework in section 4 and explain how we use it to estimate the total infections. In section 5, we evaluate the performance of MdlInfer. We then discuss future work and conclude in section 6.

## 2 Related work

### 2.1 Reported rate estimation

One of the most effective current methods to identify the reported rate in a region is through largescale serological studies [35, 16, 42]. These surveys use blood tests to identify the prevalence of antibodies against target pandemic in a large population. While serological studies can give an accurate estimation, they are expensive and are not sustainable in the long run [3]. Furthermore, it is also challenging to obtain real-time data using such studies since there are unavoidable delays between sample collection and laboratory tests [1, 16].

### 2.2 Minimum Description Length framework

MDL frameworks has been widely used for numerous optimization problems ranging from network summarization [20], causality inference [10], and failure detection in critical infrastructures [5]. They are also used in machine learning as regularization to help with model selection and avoid overfitting. [9] However, these works are built on networks and agent-based models. To the best of our knowledge, we are the first to propose an MDL-based approach on top of ODE-based epidemiological models.

## 3. Preliminaries

### 3.1 ODE-based Models

An ODE-based epidemiological model uses ordinary differential equations to describe the spread of diseases by modeling changes in populations (e.g., susceptible, infected, recovered) over time [18]. In general, the *O*_M_ has a set of parameters Θ that need to estimate from *observed data* using a so-called calibration procedure, Calibrate. In practice, the data we use for calibration can be the time series of the number of reported infections, or *D*_reported_. To estimate the number of total infections, these models often explicitly include reported rate as one of their parameters, or include multiple parameters that jointly account for it. We call it as BaseInfer in later sections for brief. There are many calibration procedures commonly used in literature, such as RMSE-based [14] or Bayesian approaches [19, 15]. BaseInfer is generally a complex, high-demensional problem, since there are multiple parameters interacting with each other. To make matters worse, there exist many possible parametrizations that show similar performance (e.g. in RMSE, likelihood) yet correspond to vastly different reported rates, and BaseInfer cannot select between these competing parametrizations in a principled way.

### 3.2 Two-part sender-receiver MDL framework

In this work, we use this framework to identify the total infections. The conceptual goal of the framework is to transmit the Data from the possession of the hypothetical sender *S* to the hypothetical receiver *R*. We assume the sender does this by first sending a Model and then sending the Data under this Model. In this MDL framework, we want to minimize the number of bits for this process. We do this by identifying the Model that encodes the Data such that the total number of bits needed to encode both the Model and the Data is minimized. Hence, our cost function in the total number of bits needed is composed of two parts: (i) model cost *L*(Model): The cost in bits of encoding the Model and (ii) data cost *L*(Data | Model): The cost in bits of encoding the Data given the Model. Intuitively, the idea is that a good Model will lead to a fewer number of bits needed to encode both Model and Data. The general MDL optimization problem can be formulated as follows: Given the Data, *L*(Model), and *L*(Data | Model), find Model), * such that

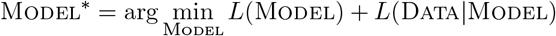

## 4 MdlInfer

### 4.1 MDL formulation

In our situation, the Data is the reported infections *D*_reported_, which is the only real-world data given to us. As for the Model, intuitively it should be 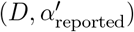 since our goal is to find the total infections *D* with the corresponding reported rate 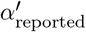.Note that as two-part MDL (and MDL in general) does not assume the nature of the Data or the Model, our MdlInfer can be applied to any ODE model. Next, we give more details how to formulate our problem of estimating total infections *D*. We also list the notations in Table 1

**Table 1:** List of notations.

| Notation | Description |
| --- | --- |
| $O_M$ | ODE model |
| $\Theta$ | ODE model parameters to infer |
| $D_{\text{reported}}$ | Reported infections |
| $\alpha_{\text{reported}}$ | Reported rate |
| BASEINFER | Baseline ODE calibration procedure<br>(calibrated on only $D_{\text{reported}}$) |
| $\hat{\Theta}$ | Parametrization estimated by BASEINFER |
| $\hat{\alpha}_{\text{reported}}$ | Reported rate in $\hat{\Theta}$ |
| $D_{\text{reported}}(\hat{\Theta})$ | ODE simulated reported infections using $\hat{\Theta}$ |
| $D(\hat{\Theta})$ | ODE simulated total infections using $\hat{\Theta}$ |
| MDLINFER | Our framework |
| $D$ | Candidate total infections in MDLINFER |
| $\Theta'$ | Parametrization estimated by MDLINFER<br>when calibrating on both $D$ and $D_{\text{reported}}$ |
| $\alpha'_{\text{reported}}$ | Reported rate in $\Theta'$ |
| $D_{\text{reported}}(\Theta')$ | ODE simulated reported infections using $\Theta'$ |
| $D(\Theta')$ | ODE simulated total infections using $\Theta'$ |

#### 4.1.1 Model space

As described above, our Model is intuitively 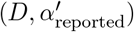.Note that reported rate is actually one of the parameters for the ODE model *O*_M_, we choose to include its corresponding parametrization 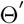 into Model. We further choose to add 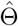 estimated by BaseInfer, making our Model to be 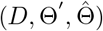.With 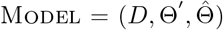,our Model space will be all possible daily sequences for *D* and all possible parametrizations for Θ′ and 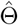.The MDL framework will search in this space to find the Model *. We also discuss other alternative Models and why 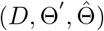 is better in Appendix.

#### 4.1.2 Model cost

With 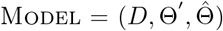,we conceptualize the model cost by imagining that the sender *S* will send the 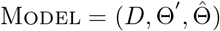 to the receiver *R* in three parts: (i) first send the 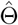 by encoding 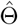 directly (ii) next send the Θ′ given 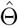 by encoding 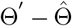 and (iii) then send *D* given Θ′ and 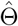 by encoding 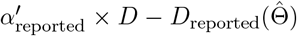.Intuitively, both 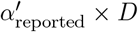 and 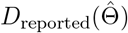 should be close to *D*_reported_, and the receiver could recover the *D* using 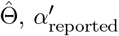,and 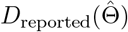 as they have already been sent. We term the model cost as 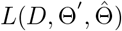 with three components: 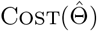,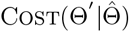, and 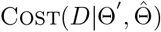,Hence

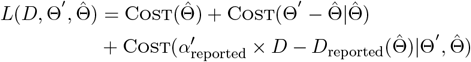

Here, the Cost(*·*) function gives the total number of bits we need to spend in encoding each term. The details of the encoding method can be found in the Appendix.

#### 4.1.3 Data cost

We need to send the Data = *D*_reported_ next given the Model. Given 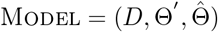,we send Data by encoding 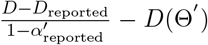.Intuitively, *D* − *D* _reported_ corresponds to the unreported infections, and 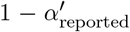 is the unreported rate. Therefore, 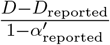 should be close to the total infections *D* and *D*(Θ′). The receiver could also recover the *D*_reported_ using 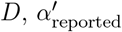,and *D*(Θ′) as they have already been sent. We term data cost as 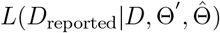 and formulate it as follows

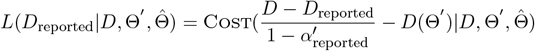

#### 4.1.4 Total cost

With 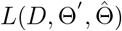 and 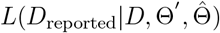 above, the total cost 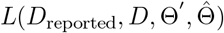 will be:

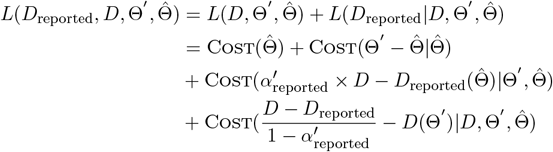

### 4.2 Problem statement

Note that our main objective is to estimate the total infections *D*. With 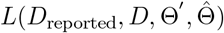,we can state the problem as: Given the time sequence *D*_reported_, epidemiological model *O*_M_, and a calibration procedure Calibrate, find *D*\* that minimizes the MDL total cost i.e.

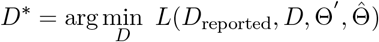

### 4.3 Algorithm

Next, we will present our algorithm to solve the problem in section 4.2. Note that directly searching *D*\* naively is intractable since *D*\* is a daily sequence not a scalar. Instead, we propose first finding a “good enough” reported rate 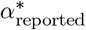 quickly with the constraint 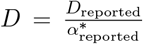 to reduce the search space. Then with this 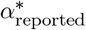,we can search for the optimal *D*\*. Hence we propose a two-step algorithm: (i) do a linear search to find a good reported rate 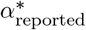 (ii) given the 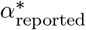 found above, use an optimization method to find the *D*\* that minimizes 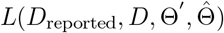 with 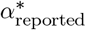 constraints. The pseudo-code is given in Algorithm 1.

#### Algorithm 1 MdlInfer

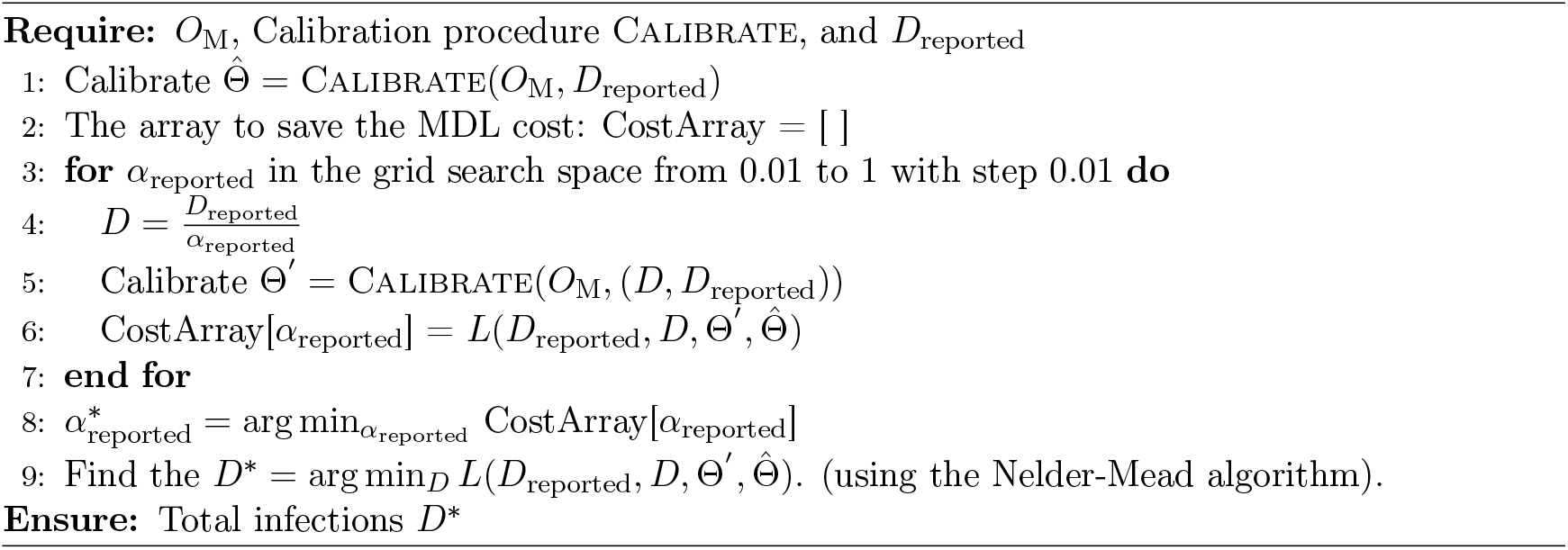

## 5 Experiments

In this section, we will answer the following research questions

- **Question 1:** Can MdlInfer estimate the total infections accurately than BaseInfer and fit the large-scale serological studies [35, 16, 42]?
- **Question 2:** Can MdlInfer fit the reported infection *D*_reported_ and forecast future infections accurately?
- **Question 3:** How does MdlInfer captures the trends of symptomatic rate?
- **Question 4:** How can MdlInfer help to evaluate the non-pharmaceutical interventions?

### 5.1 Setup

#### 5.1.1 Dataset

We choose 8 regions and periods based on the severity of the outbreak and the availability of serological studies and symptomatic surveillance data. The serological studies dataset consists of the point and 95% confidence interval estimates of the prevalence of antibodies to SARS-CoV-2 in these locations every 3–4 weeks from March to July 2020 [16, 1]. The symptomatic surveillance dataset consists of point estimate Rate_Symp_ and standard error of the COVID-related symptomatic rate starting from April 6, 2020 [29, 33]. The reported infections are from New York Times [2], which consists of the daily time sequence of reported COVID-19 infections *D*_reported_ and the mortality *D*_mortality_ (cumulative values) for each county in the US starting from January 21, 2020. In each region, we divide the timeline into two time periods: (i) observed period, when only the number of reported infections are available, and both BaseInfer and MdlInfer are used to learn the baseline parametrization (BaseParam) 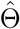 and MDL parametrization (MdlParam) 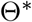,and (ii) forecast period, where we evaluate the forecasts generated by the parametrizations learned in the observed period. To handle the time-varying reported rates, we divide the observed period into multiple sub-periods and learn different reported rates for each sub-period separately. Our data and code have been deposited in https://github.com/AdityaLab/MDL-ODE-Missing, which can be run on other datasets. A demo is also deposited there.

#### 5.1.2 ODE model

We compare MdlInfer and BaseInfer using two different ODE-based epidemiological models: SAPHIRE [15] and SEIR + HD [19] as *O*_M_. Following their literature [15, 19], we use Markov Chain Monte Carlo (MCMC) as the calibration procedure Calibrate for SAPHIRE and iterated filtering (IF) for SEIR + HD, both of with are Bayesian approaches[17]. Both these epidemiological models have previously been shown to perform well in fitting reported infections and provided insight that was beneficial for the COVID-19 response.

#### 5.1.3 Metrics

To quantify the performance gap between the two approaches, we use the root mean squared error (RMSE) following the previous work [30, 31, 13, 12] for evaluation. To further demonstrate the performance, we further compute the ratio *ρ* as the fraction of the RMSE errors of BaseInfer over MdlInfer. Specifically, when the ratio is greater than 1, it implies that the MdlInfer is performing *ρ* times better than BaseInfer.

### 5.2 Q1: Estimating total infections

Here, we use the point estimates of the total infections calculated from serological studies as the ground truth (black dots shown in Figure 2). We call it SeroStudy_Tinf_. We also plot MdlInfer’s estimation of total infections, MdlParam_Tinf_, in the same figure (red curve). To compare the performance of MdlInfer and BaseInfer with SeroStudy_Tinf_, we use the cumulative value of estimated total infections. Note that values from the serological studies are not directly comparable with the total infections because of the lag between antibodies becoming detectable and infections being reported [1, 16]. In Figure 2, we have already accounted for this lag following CDC study guidelines [1, 16] (See Methods section for details). The vertical black lines shows a 95% confidence interval for SeroStudy_Tinf_. The blue curve represents total infections estimated by BaseInfer, BaseParam_Tinf_. As seen in the figure, MdlParam_Tinf_ falls within the confidence interval of the estimates given by serological studies. Significantly, in Figure 2**B** and Figure 2**F** for South Florida, BaseInfer for SAPHIRE model [15] overestimates the total infections, while for SEIR + HD model underestimates the total infections. However, MdlInfer consistently estimates the total infections correctly. This observation shows that as needed, MdlParam_Tinf_ can improve upon the BaseParam_Tinf_ in either direction (i.e., by increasing or decreasing the total infections). Note that the MdlParam_Tinf_ curves from both models are closer to the SeroStudy_Tinf_ even when the BaseParam_Tinf_ curves are different. The results of better accuracy in spite of various geographical regions and time periods show that MdlInfer is consistently able to estimate total infections more accurately.

**Figure 2.**
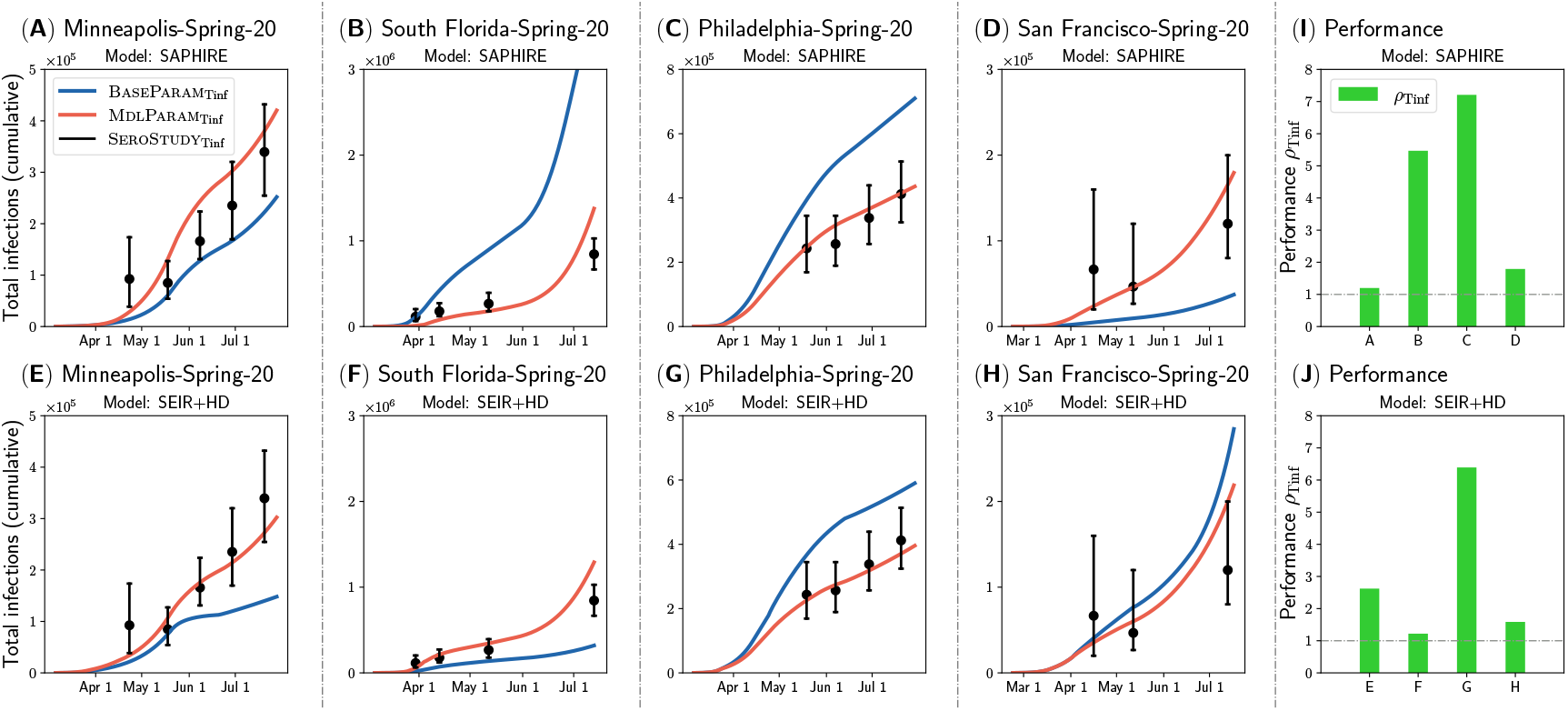
MdlInfer (red) gives a closer estimation of total infections to serological studies (black) than BaseInfer (blue) on various geographical regions and time periods. Note that both approaches try to fit the serological studies without being informed with them. (**A**)-(**H**) The red and blue curves represent MdlInfer’s estimation of total infections, MdlParam_Tinf_, and BaseInfer’s estimation of total infections, BaseParam_Tinf_, respectively. The black point estimates and confidence intervals represent the total infections estimated by serological studies [1, 16], SeroStudy_Tinf_. (**A**)-(**D**) use SAPHIRE model and (**E**)-(**H**) use SEIR + HD model. (**I**)-(**J**) The performance metric, *ρ*_Tinf_, comparing MdlParam_Tinf_ against BaseParam_Tinf_ in fitting serological studies is shown for each region. (**I**) is for SAPHIRE model in (**A**)-(**D**), and (**J**) is for SEIR + HD model in (**E**)-(**H**). Here, the values of *ρ*_Tinf_ are 1.20, 5.47, 7.21, and 1.79 in (**I**), and 2.62, 1.22, 6.39, and 1.58 in (**J**). Note that *ρ*_Tinf_ larger than 1 means that MdlParam_Tinf_ is closer to SeroStudy_Tinf_ than BaseParam_Tinf_. We show more experiments in the Appendix.

In Figure 2**I** and Figure 2**J**, we plot 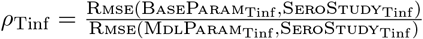. Overall, the *ρ*_Tinf_ values are greater than 1 in Figure 2**I** and Figure 2**J**, which indicates that MdlInfer performs better than BaseInfer. Note that even when the value of *ρ*_Tinf_ is 1.20 for Figure 2**A**, the improvement made by MdlParam_Tinf_ over BaseParam_Tinf_ in terms of RMSE is about 12091. Hence, one can conclude that MdlInfer is indeed superior to BaseInfer, when it comes to estimating total infections. We show more experiments in the Appendix.

### 5.3 Q2: Estimating reported infections

Here, we first use the observed period to learn the parametrizations. We then *forecast* the future reported infections (i.e., forecast periods), which were *not* accessible to the model while training. The results are summarized in Figure 3. In Figure 3**A** to Figure 3**H**, the vertical grey dash line divides the observed and forecast period. The black plus symbols represent reported infections collected by the New York Times, NYT-Rinf. The red curve represents MdlInfer’s estimation of reported infections, MdlParam_Rinf_. Similarly, the blue curve represents BaseInfer’s estimation of reported infections, BaseParam_Rinf_. Note that the curves to the right of the vertical grey line are future predictions. As seen in Figure 3, MdlParam_Rinf_ aligns more closely with NYT-Rinf than BaseParam_Rinf_, indicating the superiority of MdlInfer in fitting and forecasting reported infections.

**Figure 3.**
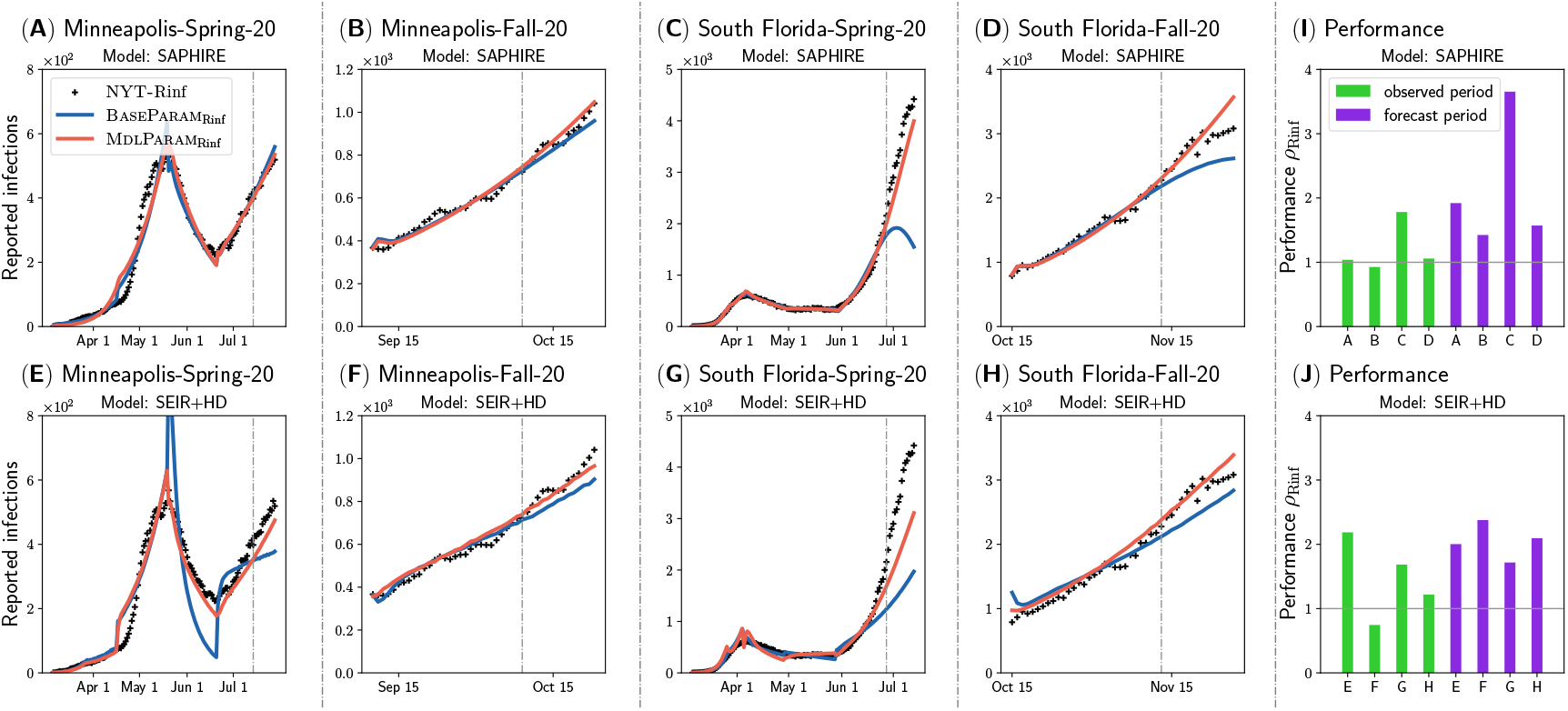
MdlInfer (red) gives a closer estimation of reported infections (black) than BaseInfer (blue) on various geographical regions and time periods. We use the reported infections in the observed period as inputs and try to forecast the future reported infections (forecast period). (**A**)-(**H**) The vertical grey dash line divides the observed period (left) and forecast period (right). The red and blue curves represent MdlInfer’s estimation of reported infections, MdlParam_Rinf_, and BaseInfer’s estimation of reported infections, BaseParam_Rinf_, respectively. The black plus symbols represent the reported infections collected by the New York Times (NYT-Rinf). (**A**)-(**D**) use SAPHIRE model and (**E**)-(**H**) use SEIR + HD model. (**I**)-(**J**) The performance metric, *ρ*_Rinf_, comparing MdlParam_Rinf_ against BaseParam_Rinf_ in fitting reported infections is shown for each region. (**I**) is for SAPHIRE model in (**A**)-(**D**), and (**J**) is for SEIR + HD model in (**E**)-(**H**). Note that *ρ*_Rinf_ larger than 1 means that MdlParam_Rinf_ is closer to NYT-Rinf than BaseParam_Rinf_. We show more experiments in the Appendix.

We use a similar performance metric 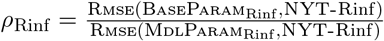 to compare MdlParam_Rinf_ against BaseParam_Rinf_ in a manner similar to *ρ*_Tinf_. In Figure 3**I** and Figure 3**J**, we plot the *ρ*_Rinf_ for the observed and forecast period. In both periods, we notice that the *ρ*_Rinf_ is close to or greater than 1. This further shows that MdlInfer has a better or at least closer fit for reported infections than BaseInfer. Additionally, the *ρ*_Rinf_ for the forecast period is even greater than *ρ*_Rinf_ for the observed period, which shows that MdlInfer performs even better than BaseInfer while forecasting.

Note that Figure 3**A, C, E, G** correspond to the early state of the COVID-19 epidemic in spring and summer 2020, and Figure 3**B, D, F, H** correspond to fall 2020. We can see that MdlInfer performs well in estimating temporal patterns at different stages of the COVID-19 epidemic. We show more experiments in the Appendix.

### 5.4 Q3: Estimating symptomatic rate trends

We validate this observation using Facebook’s symptomatic surveillance dataset [29]. We plot MdlInfer’s and BaseInfer’s estimated symptomatic rate over time and overlay the estimates and standard error from the symptomatic surveillance data in Figure 4. The red and blue curves are MdlInfer’s and BaseInfer’s estimation of symptomatic rates, MdlParam_Symp_ and BaseParam_Symp_ respectively. Note that SAPHIRE model does not contain states corresponding to the symptomatic infections. Therefore, we only focus on SEIR + HD model. We compare the trends of the MdlParam_Symp_ and BaseParam_Symp_ with the symptomatic surveillance results. We focus on trends rather than actual values because the symptomatic rate numbers could be biased [29] (see Methods section for a detailed discussion) and therefore cannot be compared directly with model outputs like what we have done for serological studies. As seen in Figure 4, MdlParam_Symp_ captures the trends of the surveyed symptomatic rate Rate_Symp_ (black plus symbols) better than BaseParam_Symp_. We show more experiments in the Appendix.

**Figure 4.**
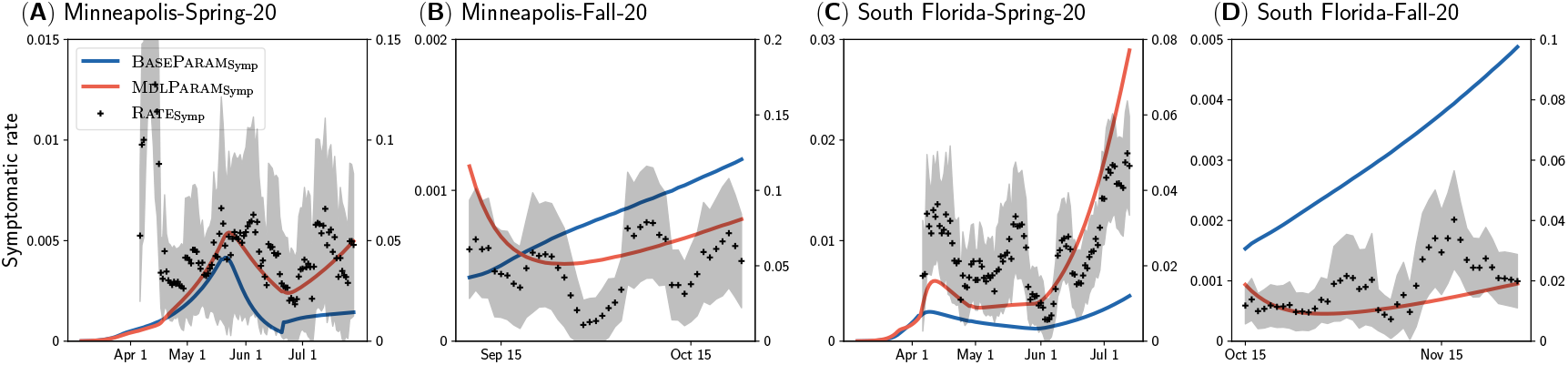
MdlInfer (red) gives a closer estimation of the trends of symptomatic rate (black) than BaseInfer (blue) on various geographical regions and time periods. (**A**)-(**D**) The red and blue curves represent MdlInfer’s estimation of symptomatic rate, MdlParam_Symp_, and BaseInfer’s estimation of symptomatic rate, BaseParam_Symp_, respectively. They use the y-scale on the left. The black points and the shaded regions are the point estimate with standard error for Rate_Symp_ (the COVID-related symptomatic rates derived from the symptomatic surveillance dataset [29, 33]). They use the y-scale on the right. Note that we focus on trends instead of the exact numbers, hence MdlParam_Symp_/BaseParam_Symp_, and Rate_Symp_ may scale differently. We show more experiments in the Appendix.

To summarize, these three sets of experiments in section 5.2 to section 5.4 together demonstrate that BaseInfer fail to accurately estimate the total infections including unreported ones. On the other hand, MdlInfer estimates total infections closer to those estimated by serological studies and better fits reported infections and symptomatic rate trends.

### 5.5 Q4: Evaluate the effect of non-pharmaceutical Interventions

We have already shown that MdlInfer is able to estimate the number of total infections accurately. In the following three observations, we show that such accurate estimations are important for evaluating the effect of non-pharmaceutical interventions.

#### 5.5.1 Non-pharmaceutical interventions on asymptomatic and presymptomatic infections are essential to control the COVID-19 epidemic

Our simulations show that non-pharmaceutical interventions on asymptomatic and presymptomatic infections are essential to control COVID-19. Here, we plot the simulated reported infections of MdlParam in Figure 5**A** (red curve). We then repeat the simulation of reported infections for 5 different scenarios: (i) isolate just the reported infections, (ii) isolate just the symptomatic infections, and isolate symptomatic infections in addition to (iii) 25%, (iv) 50%, and (v) 75% of both asymptomatic and presymptomatic infections. In our setup, we assume that the infectivity reduces by half when a person is isolated. As seen in Figure 5**A**, when only the reported infections are isolated, there is almost no change in the “future” reported infections. However, when we isolate both the reported and symptomatic infections, the reported infections decreases significantly. Even here, the reported infections are still not in decreasing trend. On the other hand, non-pharmaceutical interventions for some fraction of asymptomatic and presymptomatic infections make reported infections decrease. Thus, we can conclude that NPIs on asymptomatic infections are essential in controlling the COVID-19 epidemic.

**Figure 5.**
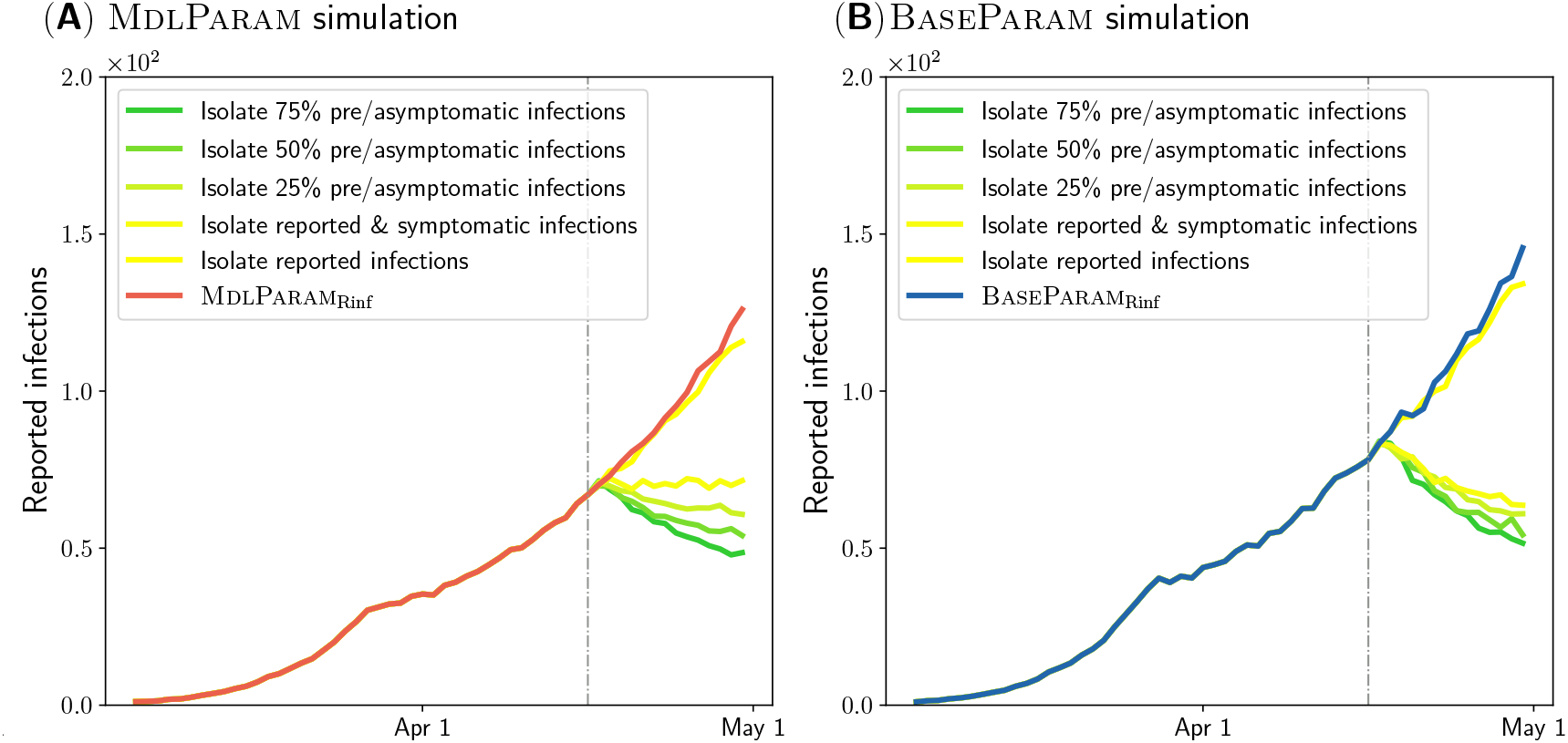
(**A**) MdlInfer reveals that non-pharmaceutical interventions (NPI) on asymptomatic and presymptomatic infections are essential to control the COVID-19 epidemic. Here, the red curve and other five curves represent the MdlInfer’s estimation of reported infections for no NPI scenario and 5 different NPI scenarios described in the Results section. The vertical grey dash line divides the observed period (left) and forecast period (right). (**B**) Inaccurate estimation by BaseInfer may lead to wrong NPI conclusions. The blue curve and other five curves represent the BaseInfer’s estimation of reported infections for no NPI scenario and the same 5 scenarios in (**B**).

#### 5.5.2 Accuracy of non-pharmaceutical intervention simulations relies on the good estimation of parametrization

Next, we also plot the simulated reported infections generated by BaseInfer in Figure 5**B** (blue curve). As seen in the figure, based on BaseInfer, we can infer that only non-pharmaceutical interventions on symptomatic infections are enough to control the COVID-19 epidemic. However, this has been proven to be incorrect by prior studies and real-world observations [24]. Therefore, we can conclude that the accuracy of non-pharmaceutical intervention simulation relies on the quality of the learned parametrization.

## 6 Conclusion

This study proposes MdlInfer, a data-driven model selection approach that automatically estimates the number of total infections using epidemiological models. Our approach leverages the information theoretic Minimum Description Length (MDL) principle and addresses several gaps in current practice including the long-term infeasibility of serological studies [16], and ad-hoc assumptions in epidemiological models [19, 23, 26, 15]. Overall, MdlInfer is a robust data-driven method to accurately estimate total infections, which will help data scientists, epidemiologists, and policy-makers to further improve existing ODE-based epidemiological models, make accurate forecasts, and combat future pandemics. More generally, MdlInfer opens up a new line of research in epidemic modeling using information theory.

## Data Availability

All data produced in the present work are contained in the manuscript.

https://www.nytimes.com/interactive/2020/us/coronavirus-us-cases.html

https://delphi.cmu.edu/covidcast/surveys/

## Acknowledgements

This paper was partially supported by the NSF (Expeditions CCF-1918770 and CCF-1918656, CAREER IIS-2028586, RAPID IIS-2027862, Medium IIS-1955883, Medium IIS-2106961, Medium IIS-2403240, IIS-1931628, IIS-1955797, IIS-2027848, IIS-2331315), NIH 2R01GM109718, CDC MInD program U01CK000589, ORNL, Dolby faculty research award, and funds/computing resources from Georgia Tech and GTRI. B. A. was in part supported by the CDC MInD-Healthcare U01CK000531-Supplement. A.V.’s work is also supported in part by grants from the UVA Global Infectious Diseases Institute (GIDI).

