## Appendix for "Accurately Estimating Unreported Infections using Information Theory"

### 1 Data

#### 1.1 New York Times reported infections [2]

This dataset (NYT-Rinf) consists of the time sequence of reported infections $D_{\text{reported}}$ and reported mortality $D_{\text{mortality}}$ in each county across the U.S. since the beginning of the COVID-19 pandemic (January 21, 2020) to current. For each county, the NYT-Rinf dataset provides the date, FIPS code, and the cumulative values of reported infections and mortality. Here, we use the averaged counts over 14 days to eliminate noise.

#### 1.2 Serological studies [6, 1]

This dataset consists of the point estimate and 95% confidence interval of the prevalence of antibodies to SARS-CoV-2 in 10 US locations every 3-4 weeks during March to July 2020. The serological studies use the blood specimens collected from population. For each location, CDC collects 1800 samples approximately every 3-4 weeks. Using the prevalence of the antibodies and the population, we can compute the estimated total infections and 95% confidence interval in the location. However, we cannot compare this number with the epidemiological model estimated total infection numbers directly as mentioned in the main article Methods section. We account for this problem by comparing the serological studies numbers with the estimated total infections of 7 days prior to the first day of specimen collection period (as suggested by the CDC serological studies work [6]).

#### 1.3 Symptomatic surveillance [9]

This dataset comes from Facebook's symptomatic survey [9]. The survey started on April 6, 2020 to current. As of January 28, 2021, there were a total of 16,398,000 participants, with the average daily participants number of 55,000. The survey asks a series of questions designed to help researchers understand the spread of COVID-19 and its effect on people in the United States. For the signal, they estimate the percentage of self-reported COVID-19 symptoms in population defined as fever along with either cough, shortness of breath, or difficulty breathing [9]. The dataset also includes weighted version which accounts for the differences between Facebook users and the United States population. In the experiments, we contrast the symptomatic rate trends inferred by our approach against the weighted data from the survey.

### 2 ODE Model

#### 2.1 SAPHIRE Model

We use the SAPHIRE model [7] as one epidemiological model $O_{\text{M}}$ in our experiments. The compartmental diagram of SAPHIRE model is shown in Figure S1. The SAPHIRE model has 9 different parameters. Note that only two parameters are calibrated, while the rest are fixed.

In this article, we expect the epidemiological model to calibrate on both reported infections $D_{\text{reported}}$ and candidate unreported infections $D_{\text{unreported}}$. We compute the newly reported infections and unreported infections as follows:

1. New reported infections= $\frac{\alpha P}{D_p}$: $\frac{P}{D_p}$ represents the number of new infections from presymptomatic infections every day in $O_{\text{M}}$. Here, we assume $\alpha$ proportion of new infections every day will be that day's new reported infections.

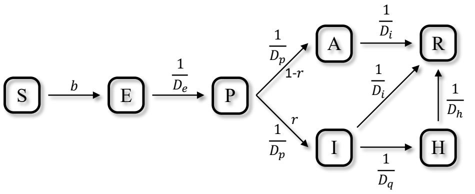


Figure S1: Compartmental diagram of SAPHIRE model [5].

2. New unreported infections $= \frac{(1-\alpha)P}{D_p}$: Then, the $1-\alpha$ proportion of new infections every day will be that day's new unreported infections.

#### 2.2 SEIR+HD Model

We also use the SEIR+HD model [7] as another epidemiological model $O_{\mathrm{M}}$ in our experiments. The compartmental diagram of SEIR+HD model is shown in Figure S2. The SEIR+HD model has of 21 different parameters. Note that only three parameters are calibrated, while the rest are fixed.

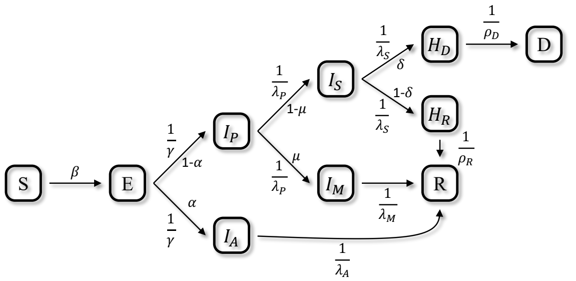


Figure S2: Compartmental diagram of SEIR+HD model [5].

Similarly to SAPHIRE model, we still expect the epidemiological model to calibrate on reported infections $D_{\text{reported}}$ and candidate unreported infections $D_{\text{unreported}}$. Hence we extend its calibration procedure to infer two more parameters: $\alpha$ and $\alpha_1$ (proportion of new symptomatic infections that are reported). We compute the newly reported infections and unreported infections as follows:

1. New reported infections$= \alpha_1 \times (N_{I_P I_S} + N_{I_P I_M})$:

   $I_{\text{new sympt}} = N_{I_P I_S} + N_{I_P I_M}$ represents the number of new symptomatic infections every day in $O_{\mathrm{M}}$. Here, we assume $\alpha_1$ proportion of new symptomatic infections every day will be that day's new reported infections.

2. New unreported infections $= (1-\alpha_1) \times (N_{I_P I_S} + N_{I_P I_M}) + N_{E I_A}$:

   Then, the $1-\alpha_1$ proportion of new symptomatic infections every day and new asymptomatic infections every day will be that day's new unreported infections.

##### 2.3 Baseline Parametrization

By calibrating $O_{\mathrm{M}}$ on $D_{\mathrm{reported}}$, we get the baseline paramterization $\mathbf{p}$:

$$\mathbf{p} = \textsc{Calibrate}(O_{\mathrm{M}}, \{D_{\mathrm{reported}}, \mathrm{others}\})$$

By running the epidemiological model with $\mathbf{p}$, $O_{\mathrm{M}}$ will output the estimated reported infections $D_{\mathrm{reported}}(\mathbf{p})$, estimated unreported infections $D_{\mathrm{unreported}}(\mathbf{p})$, and estimated total infections $D(\mathbf{p}) = D_{\mathrm{reported}}(\mathbf{p}) + D_{\mathrm{unreported}}(\mathbf{p})$. We can also calculate the reported rate $\alpha_{\mathrm{reported}}$ as follows:

$$\alpha_{\mathrm{reported}} = \frac{\sum D_{\mathrm{reported}}(\mathbf{p})}{\sum D(\mathbf{p})}$$

Here, we sum over the daily sequence $D_{\mathrm{reported}}(\mathbf{p})$ and $D(\mathbf{p})$ to calculate a scalar as the reported rate for MDL formulation.

##### 2.4 MdlInfer Parametrization

Similarly by calibrating $O_{\mathrm{M}}$ on $D_{\mathrm{reported}}$ and $D_{\mathrm{unreported}}$, we get the candidate paramterization **p'**:

$$\mathbf{p'} = \textsc{Calibrate}(O_{\mathrm{M}}, \{D_{\mathrm{reported}}, D_{\mathrm{unreported}}, \mathrm{others}\})$$

By running the epidemiological model with **p'**, $O_{\mathrm{M}}$ will output the estimated reported infections $D_{\mathrm{reported}}(\mathbf{p'})$, estimated unreported infections $D_{\mathrm{unreported}}(\mathbf{p'})$, and estimated total infections $D(\mathbf{p'}) = D_{\mathrm{reported}}(\mathbf{p'}) + D_{\mathrm{unreported}}(\mathbf{p'})$. Similarly, we can calculate the reported rate $\alpha'_{\mathrm{reported}}$ as follows:

$$\alpha'_{\mathrm{reported}} = \frac{\sum D_{\mathrm{reported}}(\mathbf{p'})}{\sum D(\mathbf{p'})}$$

With the calibration process, **p**, and **p'** defined, we can next formalize the MDL cost.

#### 3 Methodology

##### 3.1 Sender-receiver Framework

Here, we use the two-part sender-receiver framework based on the Minimum Description Length (MDL) principle. The goal of the framework is to transmit the Data in possession of the Sender $S$ to the receiver $R$ using a Model. We do this by identifying the Model that describes the Data such that the total number of bits needed to encode both the Model and the Data is minimized. The number of bits required to encode both the Model and the Data is given by the cost function $L$, which has two components: (i) model cost $L(\textsc{Model})$: The cost in bits of encoding the Model, and (ii) data cost $L(\textsc{Data}|\textsc{Model})$: The cost in bits of encoding Data given the Model.

##### 3.2 Model Space: Other Choice

In this work, the Data is $D_{\mathrm{reported}}$. One idea for defining the Model space is to use $\mathbf{p}$. With such a Model, the receiver $R$ can easily compute first $D_{\mathrm{reported}}(\mathbf{p})$ given $\mathbf{p}$. Then the sender $S$ will only need to encode and send the difference between $D_{\mathrm{reported}}(\mathbf{p})$ and $D_{\mathrm{reported}}$ so that the receiver can recover the Data fully. However, this has the disadvantage that slightly different $\mathbf{p}$ could lead to vastly different $D_{\mathrm{reported}}(\mathbf{p})$, and so the optimization problem will become hard to solve. To account for this, we propose Model as $\textsc{Model} = (D, \mathbf{p'}, \mathbf{p})$ as described in the main article, which consists of three components.

##### 3.3 Model Cost

With the model space $\textsc{Model} = (D, \mathbf{p'}, \mathbf{p})$, the sender $S$ will send the $\textsc{Model}$ to the receiver $R$ in three parts: (i) first send $\mathbf{p}$, (ii) next send $\mathbf{p'}$ given $\mathbf{p}$, and then (iii) send $D$ given $\mathbf{p'}$ and $\mathbf{p}$. Therefore, the model cost $L(D, \mathbf{p'}, \mathbf{p})$ will also have three components

$$L(D, \mathbf{p'}, \mathbf{p}) = \textsc{Cost}(\mathbf{p}) + \textsc{Cost}(\mathbf{p'}|\mathbf{p}) + \textsc{Cost}(D|\mathbf{p'}, \mathbf{p})$$

Here, we will send the first component, $\mathbf{p}$, directly, send the second component, $\mathbf{p'}$ given $\mathbf{p}$, via sending $\mathbf{p'} - \mathbf{p}$, and send the third component, $D$ given $\mathbf{p'}$ and $\mathbf{p}$, via sending $\alpha'_{\text{reported}} \times D - D_{\text{reported}}(\mathbf{p})$. We further write the model cost as below:

$$\begin{aligned} L(D, \mathbf{p'}, \mathbf{p}) &= \textsc{Cost}(\mathbf{p}) + \textsc{Cost}(\mathbf{p'} - \mathbf{p}|\mathbf{p}) \\ &\quad + \textsc{Cost}(\alpha'_{\text{reported}} \times D - D_{\text{reported}}(\mathbf{p})|\mathbf{p'}, \mathbf{p}) \end{aligned}$$

##### 3.4 Data Cost

Give the $\textsc{Model} = (D, \mathbf{p'}, \mathbf{p})$ and model cost above, next we will send the $\textsc{Data}$ in terms of the $\textsc{Model}$. Here, the $\textsc{Data}$ is $D_{\text{reported}}$, and the data cost will have only one component:

$$L(D_{\text{reported}}|D, \mathbf{p'}, \mathbf{p}) = \textsc{Cost}(D_{\text{reported}}|D, \mathbf{p'}, \mathbf{p})$$

Here, we will send it via $\frac{D - D_{\text{reported}}}{1 - \alpha'_{\text{reported}}} - D(\mathbf{p'})$, and we further write the data cost as below:

$$L(D_{\text{reported}}|D, \mathbf{p'}, \mathbf{p}) = \textsc{Cost}(\tfrac{D - D_{\text{reported}}}{1 - \alpha'_{\text{reported}}} - D(\mathbf{p'})|D, \mathbf{p'}, \mathbf{p})$$

##### 3.5 Total Cost

The total cost is the sum of model cost $L(D, \mathbf{p'}, \mathbf{p})$ and data cost $L(D_{\text{reported}}|D, \mathbf{p'}, \mathbf{p})$:

$$\begin{aligned} L(D_{\text{reported}}, D, \mathbf{p'}, \mathbf{p}) &= L(D, \mathbf{p'}, \mathbf{p}) + L(D_{\text{reported}}|D, \mathbf{p'}, \mathbf{p}) \\ &= \textsc{Cost}(\mathbf{p}) + \textsc{Cost}(\mathbf{p'}|\mathbf{p}) \\ &+ \textsc{Cost}(D|\mathbf{p'}, \mathbf{p}) + \textsc{Cost}(D_{\text{reported}}|D, \mathbf{p'}, \mathbf{p}) \\ &= \textsc{Cost}(\mathbf{p}) + \textsc{Cost}(\mathbf{p'} - \mathbf{p}|\mathbf{p}) \\ &+ \textsc{Cost}(\alpha'_{\text{reported}} \times D - D_{\text{reported}}(\mathbf{p})|\mathbf{p'}, \mathbf{p}) \\ &+ \textsc{Cost}(\frac{D - D_{\text{reported}}}{1 - \alpha'_{\text{reported}}} - D(\mathbf{p'})|D, \mathbf{p'}, \mathbf{p}) \end{aligned}$$

##### 3.6 Cost Derivation

Next, we derive the cost for each component and give our encoding method explicitly:

1. $\textsc{Cost}(\mathbf{p})$: We represent $\mathbf{p}$ as a vector of real numbers (we describe our encoding later below).
2. $\textsc{Cost}(\mathbf{p'} - \mathbf{p}|\mathbf{p})$: We will encode the difference of two vectors as a vector of real numbers.
3. $\textsc{Cost}(\alpha'_{\text{reported}} \times D - D_{\text{reported}}(\mathbf{p})|\mathbf{p'}, \mathbf{p})$: Here, we encode the difference between the two time sequences: $\alpha'_{\text{reported}} \times D$ given $D_{\text{reported}}(\mathbf{p})$.
4. $\textsc{Cost}(\frac{D - D_{\text{reported}}}{1 - \alpha'_{\text{reported}}} - D(\mathbf{p'})|D, \mathbf{p'}, \mathbf{p})$: Again, we encode it as a difference between the two time sequences: $\frac{D_{\text{unreported}}}{1 - \alpha'_{\text{reported}}}$ given $D(\mathbf{p'})$.

Next, we describe the encoding cost of real numbers, vectors, and the difference between two time sequences.

##### 3.6.1 Encoding Integers

To encode a positive integer $n$, we encode both the binary representation of integer $n$ as well as the length of the representation $\log_2 n$. Following [8], we use the cost in bits of encoding a single integer $n$ is as follows:

$$\textsc{Cost}(n) = \log_2 c_0 + \log^*(n).$$

where $c_0 \approx 2.865$ and $\log^*(n) = \log_2 n + \log_2 \log_2 n + \cdots$ as described in [8]. There are infinite terms in $\log^*(n)$ function since after we encoded a number, we always need to encode its length as another number, which could be repeated for infinite times. Additionally, if we want to transmit an integer that can be either positive or negative, we can add another sign bit and therefore the cost in bits for integers will be

$$\textsc{Cost}(n) = \textsc{Cost}(|n|) + 1.$$

##### 3.6.2 Encoding Real Numbers

Note that most real numbers (e.g. $\pi$ or $e$) need infinite number of bits to encode. Hence, we introduce a precision threshold $\delta$. With threshold $\delta$, we approximate a real number $x$ with $x_\delta$ which satisfies $|x - x_\delta| < \delta$, and we encode $x_\delta$ instead. To encode $x_\delta$, we encode both the integer part $\lfloor x \rfloor$ as well as the fractional part $x_\delta - \lfloor x \rfloor$. Hence the cost in bits of encoding a real number $x$ is as follows:

$$\textsc{Cost}(x) = \textsc{Cost}(\lfloor x \rfloor) + \log_2 \frac{1}{\delta}$$

where $\lfloor x \rfloor$ is the floor of $x$ and therefore is a integer, whose encoding cost is $\textsc{Cost}(\lfloor x \rfloor) = \log_2 c_0 + \log^*(\lfloor x \rfloor)$. Additionally, if we want to transmit a real number that can be either positive or negative, we can add another sign bit and therefore the cost in bits for real numbers will be

$$\textsc{Cost}(x) = \textsc{Cost}(|x|) + 1$$

##### 3.6.3 Encoding Vectors

To encode a vector $\mathbf{p} = [\mathbf{p}[1], \mathbf{p}[2], \cdots, \mathbf{p}[n]]$, we encode every components one by one as real numbers. Hence the cost in bits of encoding a vector $\mathbf{p}$ is as follows:

$$\textsc{Cost}(\mathbf{p}) = \textsc{Cost}(\mathbf{p}[1]) + \textsc{Cost}(\mathbf{p}[2]) + \cdots + \textsc{Cost}(\mathbf{p}[n])$$

##### 3.6.4 Encoding The Difference between Two Time Sequences

To encode the difference $A - B = [A_{t_1} - B_{t_1}, A_{t_2} - B_{t_2}, \cdots, A_{t_n} - B_{t_n}]$ between two time sequence $A = [A_{t_1}, A_{t_2}, \cdots, A_{t_n}]$ and $B = [B_{t_1}, B_{t_2}, \cdots, B_{t_n}]$, we encode every components one by one as real numbers. Hence the cost in bits of encoding the difference is as follows:

$$\begin{aligned}\textsc{Cost}(A - B) = &\ \textsc{Cost}(A_{t_1} - B_{t_1}) + \textsc{Cost}(A_{t_2} - B_{t_2}) \\ &+ \cdots + \textsc{Cost}(A_{t_n} - B_{t_n})\end{aligned}$$

#### 3.7 Problem Statement

Now we have derived every cost involved in our problem, and we can finally state our problem as one of estimating the total infections $D$ as follows: Given the time sequence $D_{\text{reported}}$ and epidemiological model $O_{\text{M}}$, find $D^*$ that minimizes the MDL total cost:

$$D^* = \arg\min_D L(D_{\text{reported}}, D, \mathbf{p'}, \mathbf{p})$$

We will give the algorithm to find such $D^*$ as follows:

#### 3.8 Algorithms

Before presenting our algorithm to find $D^*$, we will first address the problem of searching $D^*$ directly. Note that $D^*$ is a time sequence of total infections instead of a scalar, naively searching $D^*$ directly in large search space is intractable. Hence, we propose an alternate method: First, we can find quickly a good reported rate $\alpha^*_{\text{reported}}$ since we can constrain $D = \frac{D_{\text{reported}}}{\alpha_{\text{reported}}}$ to reduce the search space. Then we can search for the optimal $D^*$ with $\alpha^*_{\text{reported}}$ from step 1 as constraints. Here, we write down our two-step search algorithm to find the $D^*$ as follows:

1. Step 1: We do a linear search to find a good reported rate $\alpha^*_{\text{reported}}$, which serves as an initialization in the second step.

2. Step 2: Given the $\alpha^*_{\text{reported}}$ found in step 1, we use the Nelder-Mead [4] optimization to find the $D^*$ that minimizes $L(D_{\text{reported}}, D, \mathbf{p'}, \mathbf{p})$ with $\alpha^*_{\text{reported}}$ constraints.

##### 3.8.1 Step 1: Find the $\alpha^*_{\text{reported}}$

In step 1, we search on $\alpha_{\text{reported}}$ to find the $\alpha^*_{\text{reported}}$ as follows:

$$\alpha^*_{\text{reported}} = \arg \min_{\alpha_{\text{reported}}} L(D_{\text{reported}}, \frac{D_{\text{reported}}}{\alpha_{\text{reported}}}, \mathbf{p'}, \mathbf{p})$$

To be more specific, in the first step of our algorithm, we do a linear search on different $\alpha_{\text{reported}} = [0.01, 0.02, 0.03, \cdots, 0.99]$ and calibrate the $O_{\text{M}}$ on $D = \frac{D_{\text{reported}}}{\alpha_{\text{reported}}}$, which means

$$\mathbf{p'} = \textsc{Calibrate}(O_{\text{M}}, \{D_{\text{reported}}, \tfrac{D_{\text{reported}}}{\alpha_{\text{reported}}} - D_{\text{reported}}, \text{others}\})$$

Then we pick the $\alpha^*_{\text{reported}}$ that corresponds to the lowest total cost $L(D_{\text{reported}}, D, \mathbf{p'}, \mathbf{p})$ as the $\alpha^*_{\text{reported}}$.

##### 3.8.2 Step 2: Find the $D^*$ given $\alpha^*_{\text{reported}}$

With $\alpha^*_{\text{reported}}$ inferred in step 1, we will next find the $D^*$ that minimizes the total cost.

$$D^* = \arg \min_{D} L(D_{\text{reported}}, D, \mathbf{p'}, \mathbf{p})$$

Since we have already found $\alpha^*_{\text{reported}}$ in step 1, we will only search the $D^*$ that satisfies

$$\sum D^* = \frac{\sum D_{\text{reported}}}{\alpha^*_{\text{reported}}}$$

To search for the optimal $D^*$, we leverage the popular Nelder-Mead search algorithm [4].

### 4 Metrics

To evaluate the performance of MdlInfer, as mentioned in the main article, we use the Root Mean Squared Error (RMSE) following previous works [10, 11, 3]. Specifically, the metrics are calculated using the following equataions following [12].

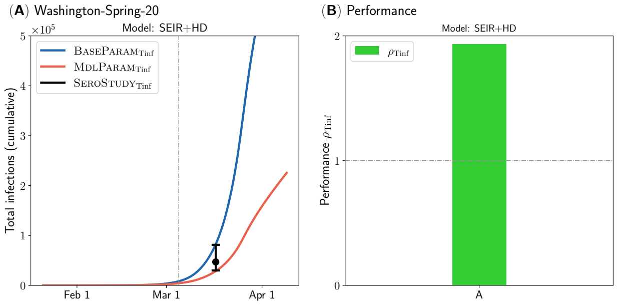


Figure S3: MDLPARAM (red) gives a closer estimation of total infections to serological studies (black) than BASEPARAM (blue). Note that the serological studies are not informed for both approaches. **a** The red and blue curves represent MDLINFER's estimation of total infections, $\text{MDLPARAM}_{\text{Tinf}}$, and baseline calibration procedure's estimation of total infections, $\text{BASEPARAM}_{\text{Tinf}}$, respectively. The black point estimates and confidence intervals represent the total infections estimated by serological studies [1, 6], or $\text{SEROSTUDY}_{\text{Tinf}}$. **b** The performance metric, $\rho_{\text{Tinf}}$, comparing $\text{MDLPARAM}_{\text{Tinf}}$ against $\text{BASEPARAM}_{\text{Tinf}}$ is shown for **a** for the SEIR+HD model. Here, the values of $\rho_{\text{Tinf}}$ is 1.93.

$$RMSE = \sqrt{\sum_t (\hat{y}_t - y_t)^2}$$

where $\hat{y}_t$ is the estimated number by either BASEINFER or MDLINFER, $y_t$ is the ground-truth value.

### 5 Experimental Setup

Here we describe our experimental setup in more detail and present results on additional testbeds. We also list the notations used in the experiments section in Table S1.

#### 5.1 Total Infections

The Results section in the main paper refers to $\text{BASEPARAM}_{\text{Tinf}}$, which represents the cumulative total infections derived from the baseline calibration procedure. It is computed as follows:

$$\text{BASEPARAM}_{\text{Tinf}} = \sum D(\mathbf{p})$$

Similarly, $\text{MDLPARAM}_{\text{Tinf}}$, which represents the cumulative total infections derived from MDLINFER, is computed as follows:

$$\text{MDLPARAM}_{\text{Tinf}} = \sum D(\mathbf{p'})$$

In Figure S3, we show additional results comparing the performance of MDLINFER and baseline calibration procedure in estimating total infections. Here, MDLINFER (red) gives a closer estimation of total infections to serological studies (black) than baseline calibration procedure (blue).

#### 5.2 Reported Infections

In Figure S4, we present additional results comparing the performance of MDLINFER and baseline calibration procedure in forecasting future infections (forecast period). Here, MDLINFER (red) gives a closer estimation of reported infections (black) than baseline calibration procedure (blue) on various geographical regions and time periods.

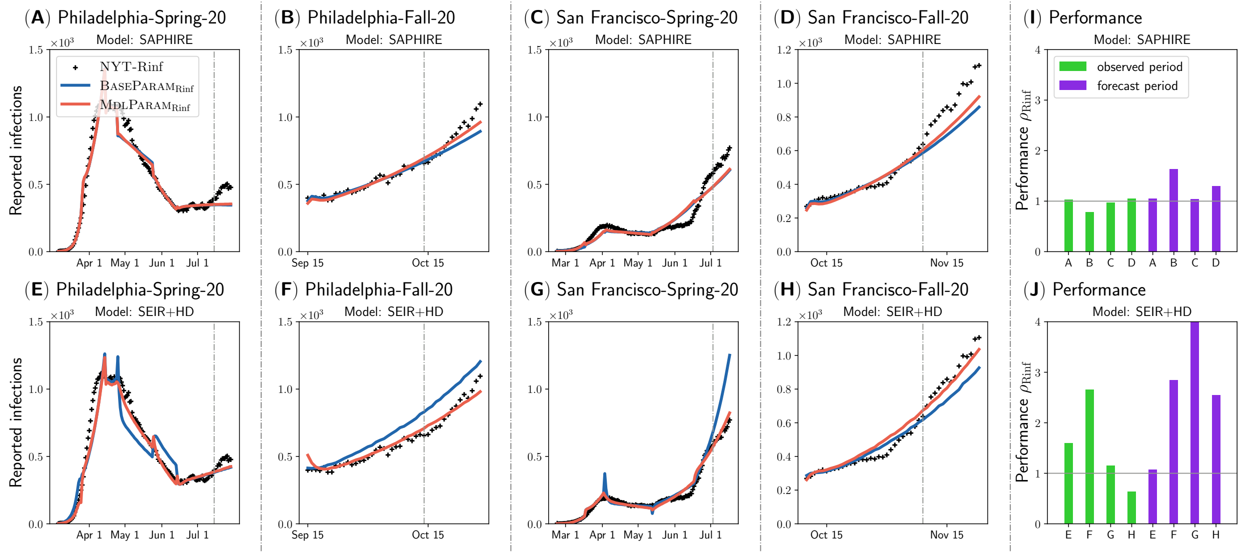


Figure S4: MdlParam (red) gives a closer estimation of reported infections (black) than BaseParam (blue) on various geographical regions and time periods. We use the reported infections in the observed period as inputs and try to forecast the future reported infections (forecast period). **a-h** The vertical grey dash line divides the observed period and forecast period. The red and blue curves represent MdlInfer's estimation of reported infections, $\text{MdlParam}_{\text{Rinf}}$, and baseline calibration procedure's estimation of reported infections, $\text{BaseParam}_{\text{Rinf}}$, respectively. The black plus symbols represent the infections reported by the New York Times (NYT-Rinf). **a-d** is for SAPHIRE model and **e-h** is for SEIR+HD model. **i-j** The performance metric, $\rho_{\text{Rinf}}$, comparing $\text{MdlParam}_{\text{Rinf}}$ against $\text{BaseParam}_{\text{Rinf}}$ is shown for **a-h** for the regions for both the SAPHIRE model in **i**, and the SEIR+HD model in **j**.

#### 5.3 Symptomatic Rate

The baseline calibration procedure and MdlInfer also estimate the symptomatic rate $\text{BaseParam}_{\text{Symp}}$ and $\text{MdlParam}_{\text{Symp}}$ respectively. We compare these against the Facebook symptomatic surveillance data $\text{Rate}_{\text{Symp}}$.

We calculate $\text{BaseParam}_{\text{Symp}}$ from $\mathbf{p}$ as follows:

$$\text{BaseParam}_{\text{Symp}} = \frac{I_S(\mathbf{p}) + I_M(\mathbf{p})}{N}$$

where $I_S(\mathbf{p})$ is the number of infections in severe symptomatic state, $I_M(\mathbf{p})$ represents the same in mild symptomatic state, and $N$ is the total population in this area.

Similarly $\text{MdlParam}_{\text{Symp}}$ is computed as follows:

$$\text{MdlParam}_{\text{Symp}} = \frac{I_S(\mathbf{p'}) + I_M(\mathbf{p'})}{N}$$

In Figure S5, we present additional results comparing MdlInfer and baseline calibration procedure in estimating trends of symptomatic rate. Here, MdlInfer (red) gives a closer estimation of symptomatic rate (black) than baseline calibration procedure (blue).

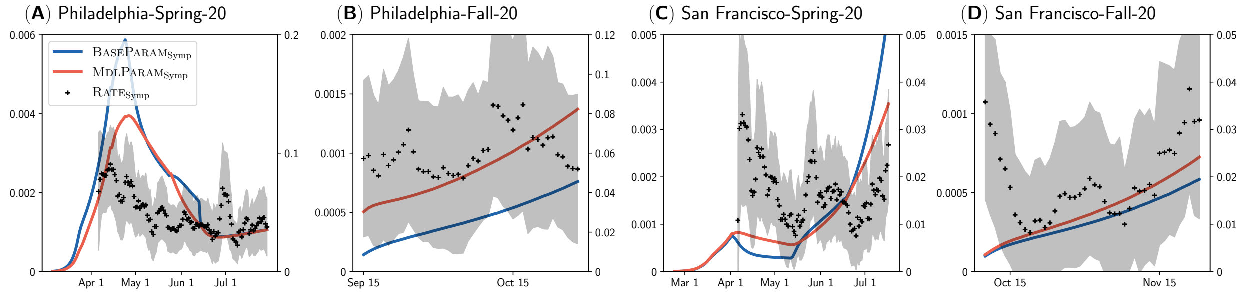


Figure S5: MdlParam (red) gives a closer estimation of symptomatic rate (black) than BaseParam (blue). The red and blue curves represent MdlInfer's estimation of symptomatic rate, $\text{MdlParam}_{\text{Symp}}$, and baseline calibration procedure's estimation of symptomatic rate, $\text{BaseParam}_{\text{Symp}}$, respectively. The black points and the shaded regions are the point estimate with standard error for $\text{Rate}_{\text{Symp}}$ (the COVID-related symptomatic rates derived from the symptomatic surveillance dataset [9, 13]).

#### 5.4 Cumulative Reported Rate

We also calculate the a dynamic reported rate from both baseline calibration procedure and MdlInfer. Note that this cumulative reported rate is different from $\alpha_{\text{reported}}$ and $\alpha'_{\text{reported}}$, which are two scalars used in MDL formulation. We calculate $\text{BaseParam}_{\text{Rate}}$ from BaseParam **p** as follows:

$$\text{BaseParam}_{\text{Rate}} = \frac{\sum \text{NYT-Rinf}}{\sum D(\mathbf{p})}$$

Similarly we calculate $\text{MdlParam}_{\text{Rate}}$ from MdlParam **p'** as follows:

$$\text{MdlParam}_{\text{Rate}} = \frac{\sum \text{NYT-Rinf}}{\sum D(\mathbf{p'})}$$

#### 5.5 Non-pharmaceutical Interventions Simulation

We also use the baseline calibration procedure and MdlInfer to perform non-pharmaceutical interventions simulation on SEIR+HD model. Here, both the baseline calibration procedure and MdlInfer are estimated on the observed period, then on the future period, we will consider the following five scenarios of isolation:

1. Isolate reported infections: We isolate the $\alpha_1$ fraction of severe symptomatic infections $I_S$ and mild symptomatic infections $I_M$.

2. Isolate both reported infections and symptomatic infections: Note that some reported infections are included in the symptomatic infections. Here, we isolate all severe symptomatic infections $I_S$ and mild symptomatic infections $I_M$.

3. Isolate 25% presymptomatic and asymptomatic infections: We isolate 25% of presymptomatic infections $I_P$, asymptomatic infections $I_A$, and all severe symptomatic infections $I_S$ and mild symptomatic infections $I_M$.

4. Isolate 50% presymptomatic and asymptomatic infections: We isolate 50% of presymptomatic infections $I_P$, asymptomatic infections $I_A$, and all severe symptomatic infections $I_S$ and mild symptomatic infections $I_M$.

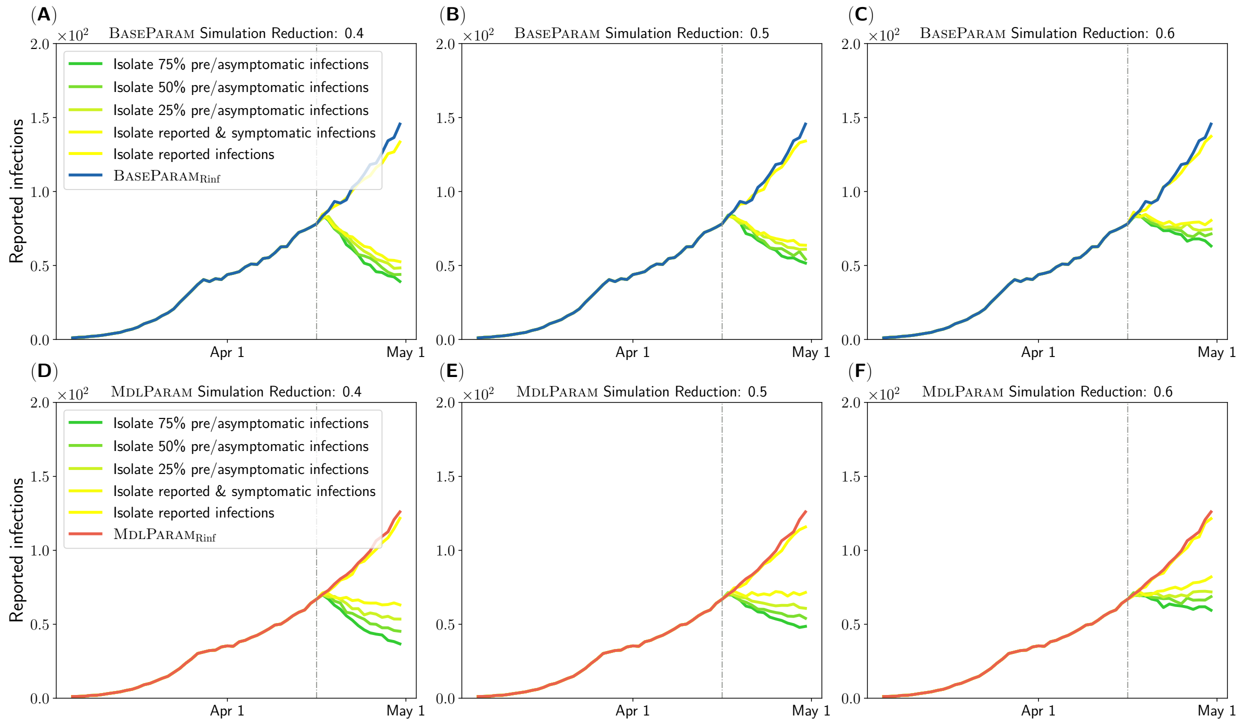


Figure S6: Our non-pharmaceutical interventions simulation results are robust. **a-c**, the vertical grey dash line divides the observed period and future period. The blue curve represents the BASEPARAM's estimation of reported infections. The other five curves represent the simulated reported infections for 5 scenarios: (i) Isolate the reported infections, (ii) symptomatic infections, symptomatic infections and (iii) 25%, (iv) 50%, (v) 75% asymptomatic and presymptomatic infections, where we reduce the infectiousness of these isolated infections to 40% in **a**, 50% in **b**, and 60% in **c** in future period. **d-f**, the vertical grey dash line divides the observed period and future period. The red curve represents the MDLPARAM's estimation of reported infections. The other five curves represent the simulated reported infections for the same 5 scenarios as in **a** to **c**. The results are for Minneapolis-Spring-20 that we shown in main article Figure 5.

5. Isolate 75% presymptomatic and asymptomatic infections: We isolate 75% of presymptomatic infections $I_P$, asymptomatic infections $I_A$, and all severe symptomatic infections $I_S$ and mild symptomatic infections $I_M$.

The infectiousness of the noes in isolated is reduces by 50%.

### 6 Sensitive Analysis

We also perform sensitivity experiments to inspect the robustness of our non-pharmaceutical interventions simulations for Minneapolis-Spring-20 in Figure S6. Here, we reduce the infectiousness of the isolated infections to 3 different values: 0.4, 0.5, and 0.6, and repeat simulations in each of the scenarios.

Our results consistently show that only isolating reported or symptomatic infections is not be enough to reduce the future reported infections. However, isolating both symptomatic infections and some fraction of asymptomatic and presymptomataic infections leads to reduction in reported infections in most settings.

Table S1: List of notations

| Symbol | Description |
|---|---|
| MdlInfer | Our Minimum Description Length (MDL) framework to estimate total infections |
| $O_{\mathrm{M}}$ | Epidemiological models used in MdlInfer |
| BaseParam | Baseline parameterization obtained via baseline calibration procedure |
| MdlParam | Optimal parametrization identified by MdlInfer |
| $\mathrm{SeroStudy}_{\mathrm{Tinf}}$ | Total infections estimated by serological studies |
| $\mathrm{BaseParam}_{\mathrm{Tinf}}$ | Total infections estimated by baseline calibration procedure |
| $\mathrm{MdlParam}_{\mathrm{Tinf}}$ | Total infections estimated by MdlInfer |
| $\rho_{\mathrm{Tinf}}$ | The performance metric comparing MdlInfer against baseline calibration procedure in estimating total infections |
| NYT-Rinf | New York Times reported infections |
| $\mathrm{BaseParam}_{\mathrm{Rinf}}$ | Reported infections estimated by baseline calibration procedure |
| $\mathrm{MdlParam}_{\mathrm{Rinf}}$ | Reported infections estimated by MdlInfer |
| $\rho_{\mathrm{Rinf}}$ | The performance metric comparing MdlInfer against baseline calibration procedure in estimating reported infections |
| $\mathrm{Rate}_{\mathrm{Symp}}$ | COVID-related symptomatic rate from symptomatic surveillance data |
| $\mathrm{BaseParam}_{\mathrm{Symp}}$ | Symptomatic rate estimated by baseline calibration procedure |
| $\mathrm{MdlParam}_{\mathrm{Symp}}$ | Symptomatic rate estimated by MdlInfer |
| $\mathrm{SeroStudy}_{\mathrm{Rate}}$ | Cumulative reported rate estimated by serological studies |
| $\mathrm{BaseParam}_{\mathrm{Rate}}$ | Cumulative reported rate estimated by baseline calibration procedure |
| $\mathrm{MdlParam}_{\mathrm{Rate}}$ | Cumulative reported rate estimated by MdlInfer |
